# Feasibility of at-home online cognitive screening prior to primary care wellness visits for older adults

**DOI:** 10.1101/2025.08.19.25333983

**Authors:** Louisa Thompson, Charles Eaton, Sarah Prieto, Molly Lawrence, Stephanie Czech, Jennifer Rosenbaum, David Anthony, Rabin Chandran, Lauren Kelly, Ivy Ngo, Arnold Goldberg, Caitlin Gillooly, Caroline Richardson, A. Rani Elwy, Richard N. Jones, Dorene M. Rentz

**Affiliations:** Department of Psychiatry & Human Behavior, Alpert Medical School of Brown University, Providence, RI, USA; Department of Family Medicine, Alpert Medical School of Brown University, Providence, RI, USA; Department of Epidemiology, School of Public Health, Brown University, Providence, RI, USA; Center for Healthcare Optimization & Implementation Research, VA Bedford Healthcare System, Bedford, MA, USA; Department of Neurology, Massachusetts General Hospital, Harvard Medical School, Boston, MA, USA; Center for Alzheimer Research & Treatment, Brigham and Women’s Hospital, Harvard Medical School, Boston, MA, USA

**Author notes:** Correspondence concerning this article should be addressed to Louisa Thompson, Department of Psychiatry, Alpert Medical School, Brown University, Providence, RI. Address: 345 Blackstone Blvd., Providence, RI 02906.

**Keywords:** Primary Care, Cognitive Screening, Alzheimer’s disease, Mild Cognitive Impairment, Digital Assessment, Early Detection

## Abstract

**BACKGROUND:** Improved methods of cognitive testing are urgently needed for primary care settings faced with a growing older adult population and higher rates of dementia. We tested the feasibility and acceptability of a novel online cognitive test that can be completed at home prior to an annual exam.

**METHODS:** 32 older adults completed testing at home on personal devices 1-4 weeks prior to an annual exam with their primary care provider. Additional cognitive screening was completed in-clinic for validation. Completion rates were examined to assess feasibility, and patient acceptability was assessed via survey.

**RESULTS:** Completion rate was 78.5% for at-home testing online. Participants reported that they generally preferred at-home over in-clinic testing. At-home test performance was moderately correlated with the standard Montreal Cognitive Assessment completed in clinic.

**CONCLUSION:** Findings support the acceptability of self-administered online cognitive screening in older adults. Completion rates were slightly below expectation, suggesting that additional support may be needed to facilitate at-home testing in routine care.

## Background

Improved detection and management of cognitive impairment is urgently needed in primary care, where approximately nine out of every ten older adults report receiving care.^1^ Recent estimates project that 42% of Americans over age 55 will eventually develop some form of dementia, with higher rates possible in women.^2^ Late onset Alzheimer’s disease (AD), the most common cause of dementia, typically has a slow onset and progression, but the majority of cases still go undetected in primary care.^3,4^ In fact, an estimated 90% of cases of mild cognitive impairment (MCI), often a precursor to AD or other dementias, go undiagnosed among Medicare beneficiaries, with higher rates missed among Black and Hispanic or Latino patients.^4,5^

Growing evidence in support of early interventions for modifiable dementia risk factors highlights the importance of identifying older adults at risk for cognitive decline as early as possible.^6,7^ At the same time, attitudes toward dementia diagnosis are also changing. A recent survey found that 79% of Americans would prefer to receive a diagnosis while symptoms are minor and do not interfere with activities of daily living.^12^ Missed and/or delayed diagnoses not only heighten the risk of disease progression but also limit opportunities for patients to adopt beneficial lifestyle changes, access treatment, and participate in care planning.^8,9^

Commercially-available plasma biomarkers that detect the earliest stages of AD have recently been FDA approved to aid in the differential diagnosis of MCI and dementia; however, use guidelines indicate that these tests should only be used in the context of confirmed cognitive decline.^10,11^ Unfortunately, existing paper-based cognitive screening tools are often insensitive for accurately detecting MCI, particularly in diverse older adults in primary care.^12^ This potentially creates a significant bottleneck in identifying appropriate candidates for biomarker testing. Several newer digital cognitive assessment tools have been shown to be feasible, sensitive to mild impairment, and highly reliable in older adults, but only a few have been validated for use in primary care.^13-15^ Moreover, given time constraints faced by primary care practitioners, there is a greater need to determine whether digital assessments could be self-administered remotely (i.e., at home) by older adults to initiate a diagnostic work up or supplement ongoing cognitive monitoring.^16^ With an estimated 88% of U.S. older adults using the internet regularly, now is the time to investigate digital alternatives to traditional testing.^17,18^

The Boston Cognitive Assessment (BOCA) is a 10-minute online test of global cognition developed for clinical use in older adults. Strong correlations between BOCA and Montreal Cognitive Assessment (MoCA) have been found in memory clinic patients and healthy controls.^19,20^ Additionally, BOCA is sensitive to cerebral amyloid accumulation, a neuroimaging biomarker of AD,^21^ making it a promising candidate for earlier detection of cognitive impairment for primary care settings. We designed and conducted the present study to evaluate the feasibility, acceptability, and preliminary validity of a remote cognitive screening intervention using the BOCA to detect cognitive impairment among older adult patients in primary care, with the following specific objectives: 1) Determine if remote testing with BOCA is feasible with minimal support based on test completion rates; 2) Survey the acceptability of digital tests among patients, and 3) Conduct a preliminary comparison of the remote digital assessment with in-clinic cognitive screening tools, including the paper-based MoCA,^22^ as a reference standard.

## Methods

### Recruitment & Enrollment

Of approximately 80 primary care providers (PCPs) contacted via email invitation to partner in the study, we engaged five across three clinics in Rhode Island. Clinic A was a hospital-affiliated community clinic with two PCPs engaged in the study, clinic B was a Federally Qualified Health Center with one PCP engaged, and clinic C was a hospital-affiliated family medicine clinic with two PCPs engaged. Using electronic medical record (EMR) searches, we identified potential participants (age range of 55-85, no dementia diagnosis) from each partnering PCP’s panel. We utilized a multipronged approach for recruitment to maximize insights for future research. Passive methods included mailings, MyChart messages (clinic A and clinic C only), and waiting room flyers. Direct recruitment included phone calls to MyChart users with preferences indicating willingness to be contacted for research, as well as calls to potential participants identified by the PCP.

Screening was conducted over the phone. Eligibility criteria required participants to be fluent in English. Exclusion criteria were existing dementia diagnosis or other neurological disease, a score of <13 on the telephone version of the MoCA^23^, current substance use disorder, acute or unstable psychiatric illness, and motor or sensory conditions that would compromise test validity based on self-report during screening. Consent was collected electronically. Ethical approval for the study was obtained from the Butler Hospital Institutional Review Board (approval #1882523). Participant compensation of $20 was provided in the form of a gift card.

### Cognitive assessment

#### Remote assessment

Remote assessment was conducted using the Boston Cognitive Assessment (BOCA) which participants completed using their own smartphone, personal computer, or tablet. To assess test-retest reliability, BOCA was completed at two timepoints approximately 1–4 weeks before their Medicare Annual Wellness Visit (AWV) or follow-up, and again within one day of the appointment (Figure 1). Participants received a link to complete BOCA via email and a reminder email or call as needed. The BOCA is a 10-minute test of global cognition with subtasks assessing orientation, immediate and delayed recall, executive function, visuospatial ability, attention, and language scored for a total of 30 points.^20^ Testing within each subtask is preceded by a practice session to help explain the task. Alternate forms are given for repeat assessment. Additional test characteristics and psychometrics have been published previously.^20^

**Figure 1.**
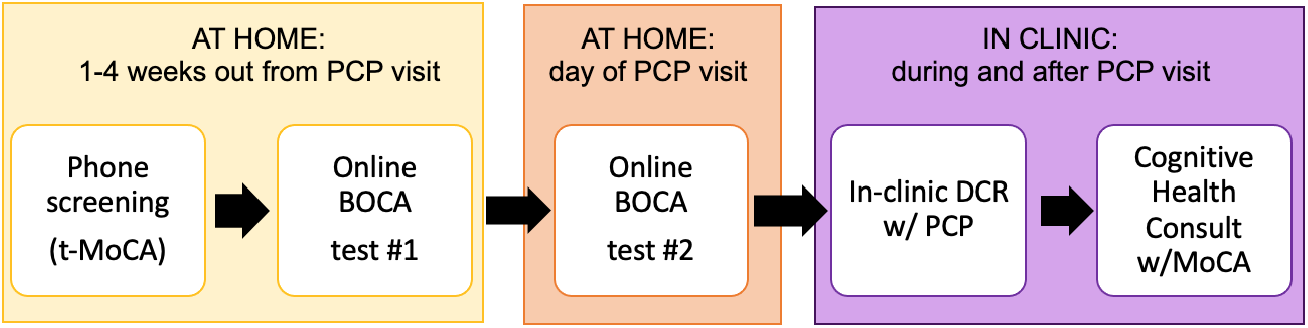
Protocol. *Note*. PCP = Primary Care Provider, t-MoCA and MoCA = Telephone version and standard version, respectively, of the Montreal Cognitive Screening Assessment, BOCA = Boston Cognitive Assessment, DCR = Digital Clock and Recall test.

**Figure 2.**
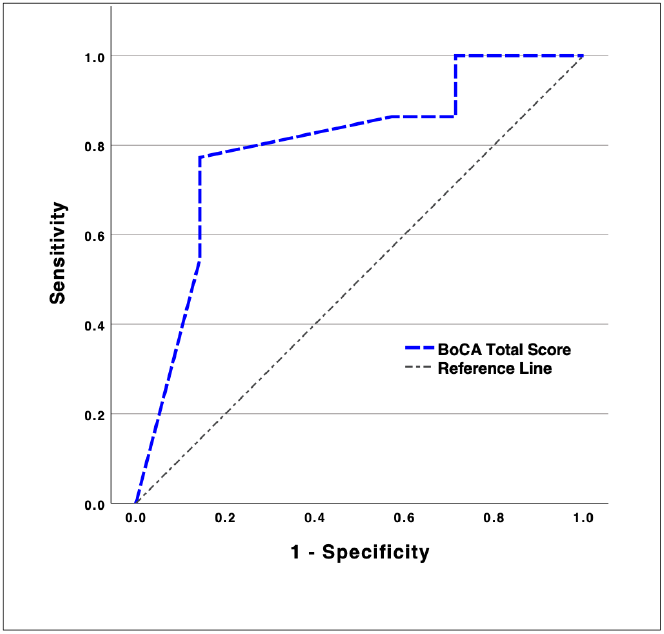
Receiver Operating Characteristic Analysis of BOCA to distinguish cognitive status. *Note*. Cognitive status of based on a MoCA score of <26 indicative of possible cognitive impairment.

#### In-clinic Assessment

At their PCP appointment, participants completed a comparison digital cognitive screening test, the Linus Health Digital Clock and Recall (DCR™), supervised by the PCP. The DCR is a 3-5-minute assessment including 3-word learning, stylus-drawn clock, and 3-word recall. A cloud-based scoring algorithm calculates a total (0–5) and sub-scores.^24-26^ As the primary reference standard for validation, participants also completed the Montreal Cognitive Assessment (MoCA) administered by research staff at the end of their appointment as part of a brain health consultation (Figure 1). MoCA results were reviewed with participants and recommendations for maintaining brain health were provided as well as recommendations for follow-up, if needed. MoCA results were scanned to the participant’s EMR with their permission. Impaired scores were communicated directly to the PCP and a follow-up referral to our memory clinic was offered.

### Survey & Interview

Participants provided feedback via a 5–10-minute online survey developed to assess acceptability, including test preference, difficulty, and usability, as well as comfort and stress associated remote versus in-clinic testing.

### Analysis

Wilcoxon rank-sum and chi-squared tests were used to examined whether there were differences in education, race, or employment status between enrolled participants and those screened but not enrolled. We set a benchmark completion rate of 80% to assess the feasibility of the remote and in-clinic digital cognitive assessment protocols. Test-retest reliability was calculated for the BOCA administered across two timepoints (1-4 weeks apart). Convergent validity was tested using Spearman’s correlations between BOCA, DCR and the MoCA. Area under the curve (AUC) analyses examined BOCA accuracy for distinguishing cognitive impairment on the MoCA using a cutoff of 26.

## Results

### Participants

A total of 48 older adults were screened for the study and 37 enrolled; however, five withdrew or were lost to follow-up before completing any study activities. Reasons for withdrawal, when given (n = 2), included not having enough time or needing to focus on other high priority health conditions. Overall attrition rate was 15.6% and the final enrolled sample size was N = 32. The sample was 56% female, 84% White and 16% Black. Mean participant age was 68 years (SD = 6.6) and mean years of education was 14.7 (SD = 2.4). Results indicated no significant differences in demographics between screened and enrolled participants.

### Protocol feasibility

#### Provider and patient engagement

Patient recruitment was limited to individuals in the care of our PCP partners. 613 mail invitations were sent to patients, yielding a 1.6% screening rate. 253 messages were sent via MyChart EMR system, yielding a 4.35% screening rate. Direct recruitment methods were more productive, with screening rates of 11.3% from 142 phone calls to MyChart users, and 30.3% from 33 direct PCP referrals. PCP direct referral was particularly useful at Clinic B, which did not use MyChart. There were no significant differences in recruitment method by participant demographics.

#### BOCA completion

Completion rate for BOCA administered at time one (T1), 2-4 weeks in advance of the PCP appointment, was 78.5%. Six participants were non-compliant with T1 BOCA despite follow-up reminders sent via email. For BOCA testing on the day of their PCP appointment (T2), completion rate was 66.7%, with six out of window completions and three non-compliant. Reasons for incompletes included time commitment (n = 4), non-compliance (n = 3), and technical difficulties (n = 2). Four participants were not assigned to complete the T2 BOCA due to insufficient time window and were not included in the completion rate calculation.

### BOCA acceptability

Twenty-two participants completed the feedback survey. 50% preferred completing cognitive testing at home, compared to 10% who preferred in-clinic testing, and another 40% who said they preferred to do both at-home and in-clinic testing. 78% reported feeling more comfortable completing cognitive testing at home, while the other 22% said they felt more comfortable in the clinic. The in-clinic testing was rated as more difficult (65%) compared to the at-home test. All reported that they would be willing to complete the at-home test again in the future. At-home testing was most often completed in the morning using a smartphone (48%) versus a computer (28%) or tablet (24%).

### Preliminary BOCA validity

BOCA test-retest reliability between the T1 and T2 timepoints was good (r = .81, 95% CI [.54 - .93], n = 17, mean time difference =15.5 days, range 4-33 days). Subsequent analyses examined only the first BOCA instance for each participant collapsed across T1 and T2. BOCA total scores were moderately correlated with the MoCA (r = .59, 95% CI [.28 - .79], p <.001) and strongly correlated with the DCR (r = .71, 95% CI [.45 - .86], p <.001). For comparison, the DCR was strongly correlated with MoCA (r = .76, 95% CI [.54 - .89], p < .001). Mean BOCA score was 27.4, SD = 3.2, while mean MoCA score was 26.5, SD =2.6. Area under the curve (AUC) analysis indicated a BOCA total score AUC =.80, 95% CI [0.60-1.0] to distinguish cognitive status (Figure 1). A cutoff score for identifying impairment has yet to be established for BOCA. Based on this preliminary ROC curve analysis, a cutoff of 24 would maximize sensitivity at .91, with a specificity of .71, which would result in a high negative predictive power but poor positive predictive power in clinical settings with similar prevalence rates of cognitive impairment (∼15%).

## Discussion

Our results indicate that unsupervised online testing with BOCA at home is an acceptable approach to cognitive screening for older adults in primary care. However, the BOCA completion rate of 78.5% did not quite meet our 80% benchmark despite the provision of reminders, suggesting that the use of BOCA or other similar tools in routine care would likely require significant support resources to facilitate completion. Future deployment of at-home, unsupervised digital tests may benefit from a designated coordinator who could enlist a family member or assist with technology support, as needed. Remote digital assessments would also benefit from integration with electronic medical records (EMR) or artificial intelligence (AI) tools for setup, reminders, and monitoring. Indeed, recent literature has also identified workflow compatibility as an important feature for digital tool selection among providers.^27^ While EMR integration is advancing and has the potential to enhance implementation, scalable protocols that do not rely on EMR are still needed for many resource-limited settings.^28^

Completing the digital cognitive testing at home was widely viewed as acceptable to patients and there were few technical challenges, suggesting that online tests completed via simple URL links are practical for remote use. Although most patients said they preferred to do testing at home and were more comfortable with testing at-home, 40% said that they would prefer to complete both at-home and in-clinic testing, suggesting patients may also see value in repeating a test in-clinic with their provider.

The BOCA demonstrated preliminary validity, showing a moderate correlation with the MoCA, which is consistent with the prior literature that was limited to secondary care settings.^21,26^ BOCA accuracy for detecting cognitive impairment was good, though there may have been ceiling effects in our small sample and specificity was limited. Results nonetheless indicate that BOCA testing prior to an annual appointment could be beneficial for first line screening, with positive results followed by confirmatory testing in-clinic. This approach could allow primary care providers to identify at-risk individuals prior to annual visits, enabling targeted follow-up while reducing in-clinic testing demands. Scalable, home-based testing could be particularly valuable given time constraints in primary care and the increasing need for early detection as disease-modifying treatments for Alzheimer’s disease emerge. Providing participants with their screening results and educational materials to support brain health awareness was a strength of this study which can be extended to larger implementation efforts.

Examining acceptability and barriers to cognitive testing, including digital and remote approaches, should be explored in future implementation studies. Interestingly, given the minimal facilitation provided for this feasibility study, providers initially chose to administer the DCR themselves despite the option to delegate to clinic support staff, citing high staff burden and turnover. These insights underscore the potential advantage of at-home testing and need to design tools and support workflows that minimize added workload for the full care team. For patient engagement, direct referrals from PCPs proved the most effective recruitment method, while passive approaches were less successful. These findings reinforce the importance of trusted provider-patient relationships to facilitate patient education and engagement in cognitive testing.

The small geographically limited sample likely constrains the generalizability of our findings. Participants were more highly educated and less racially diverse than state averages, which may have influenced performance and validity outcomes. While the MoCA cutoff of <26 was appropriate for our sample, alternative cutoffs or reference standards may be necessary for more diverse populations.^29,30^ Finally, while the MoCA offers an initial benchmark for convergent validity, more in-depth multimodal validation studies comparing novel digital tools with more extensive cognitive testing, disease biomarkers, and functional assessments are needed.

Although evaluating clinical outcomes was not a focus of this study, the involvement of patients in the study successfully facilitated several referrals for further evaluation to our memory clinic or with a neuropsychologist. Several participants with low or borderline scores were referred for further evaluation, and some were identified as having potentially treatable contributing conditions. These cases highlight the potential clinical value of integrating cognitive screening into routine care. A focus of future research will include assessing whether early detection via these tools improves dementia care outcomes.

In sum, our findings support the feasibility and acceptability of remote digital cognitive assessments in primary care but highlight the necessity of supportive infrastructure to ensure successful implementation. Continued research with larger, more diverse populations and in varied practice settings will be essential to optimize these tools for widespread adoption.

## Data Availability

All data produced in the present study are available upon reasonable request to the authors

## ACKNOWLEDGEMENTS

The research team would like to thank our digital assessment partners, including Dr. Andrey Vyshedskiy (Boston Cognitive Assessment), Dr. Alvaro Pascual-Leone (Linus Health). The Montreal Cognitive Assessment was used with permission.

## CONSENT STATEMENT

The study protocol was approved by the Butler Hospital IRB. All participants provided consent.

## CONFLICTS

All authors have nothing to disclose.

## FUNDING

This work is supported by grants from the NIA (K23AG080159, PI: Thompson) and Rhode Island Advance-CTR (IDeA-CTR, NIGMS U54GM115677).

## Notes

### Competing Interest Statement

The authors have declared no competing interest.

### Author Declarations

The IRB of Butler Hospital gave ethical approval for this work

### Summary of Updates

The previous version of the manuscript was rejected following peer review. Upon recommendation we have subsequently edited the manuscript to reduce the scope and simplify the reporting. The focus is now on the remote assessment component from the first phase of the study.

